# Cryptic spread of SARS-CoV-2 undermined targeted travel restrictions

**DOI:** 10.1101/2025.08.29.25334701

**Authors:** Praneeth Gangavarapu, Jassim Mohammed Abdo, Edyth Parker, Mark Zeller, Refugio Robles-Sikisaka, Ezra Kurzban, Sarah Perkins, Madison Schwab, Kelly Nguyen, Michelle McGraw, Karthik Ramesh, Catelyn Anderson, Narin A. Rasheed, Renas Husain Isa, Sherzad Majeed Taher, Karthik Gangavarapu, Kristian G. Andersen

## Abstract

The effectiveness of travel restrictions in mitigating the spread of novel viral variants remains understudied, particularly in regions with sparse genomic surveillance. We evaluated the efficacy of these restrictions in Western Asia, where countries sequenced less than one percent of SARS-CoV-2 cases in 2021, focusing on Iraq’s travel ban on India and the tourist travel restriction. We generated 535 SARS-CoV-2 whole-genome sequences from clinical samples collected in Duhok, Iraq, between June 2021 and March 2022, encompassing the Delta and Omicron variant waves. Using discrete Bayesian phylogeographic modeling, we reconstructed spatiotemporal viral spread patterns to determine whether travel restrictions delayed the introduction of the variants into the country, postponed community transmission onset, or altered the geographic sources of viral introductions over time. We found that the Delta variant was introduced into Iraq three months before the targeted travel ban was enacted, predominantly from Western and Southern Asia, whereas the Omicron variant was introduced despite the tourist travel restriction already being in place. We also show that both variants had established community transmission within the country despite travel restrictions, rendering these measures ineffective. Analysis of travel restrictions on a global scale reveals similar patterns of limited efficacy in mitigating the introductions of novel SARS-CoV-2 variants, challenging the usefulness of such measures.

## Introduction

The global expansion of genomic surveillance infrastructure during the COVID-19 pandemic has enabled the characterization of SARS-CoV-2 transmission dynamics at unprecedented temporal and geographic scales.^1–3^ In response to the emergence of variants of concern (VOCs) such as Delta and Omicron, several countries instituted travel restrictions, often targeting a single country where the VOC was first detected, with the stated goal of delaying the introduction of the virus.^4^ These restrictions took two primary forms, with the targeted travel ban focusing on specific countries and a tourist travel restriction limiting recreational travel. However, studies have repeatedly shown that these reactive measures implemented after the first detection of a VOC are too late to be effective and simply cause introductions of the VOC to occur via other regions.^1,2,5,6^ Consequently, studies investigating the efficacy of travel restrictions have recommended increased genomic surveillance as the more effective alternative to mitigate the spread of a VOC.^2,3,7–9^ However, the generalizability of these findings to different regions is limited by heterogeneity in mobility networks, socio-political, and economic connections to neighbors, and genomic sequencing coverage. These limitations are particularly pronounced in Western Asia, where very few studies have investigated transmission drivers of the virus.^10,11^ Among these studies, Parker et al. 2022 specifically examined Jordan’s first (driven by B.1.1.312 and B.1.36.10) and second (driven by B.1.1.7) waves and found that regional connectivity, specifically land-based travel covering short distances, disproportionately drove transmission of SARS-CoV-2 between Western Asian regions even when travel restrictions were in effect.^11^ However, the applicability of these findings remains uncertain given the increased transmissibility of the Delta and Omicron variants.^12,13^

The Delta variant (B.1.617.2 and its sublineages) was first detected in India in March 2021, though it likely emerged in mid-October 2020, and demonstrated higher replication efficiency and immune evasion than prior variants.^12,14^ The variant circulated cryptically for six months, and by May 2021, had spread to at least 97 countries and evolved into more than 120 classified sublineages.^15^ In the fall of 2021, the emergence and global expansion of the Delta variant was followed by the rapid emergence of another VOC, the Omicron variant (BA.1, its sister lineages and their sublineages). The Omicron variant was identified in 87 countries within just one month of its detection in November 2021 in southern Africa.^16^ Omicron proved particularly challenging because it demonstrated a higher binding affinity for ACE2 and higher immune evasion than the Delta variant.^13,17^ In response to the emergence of these two VOCs, Iraq enacted several travel restrictions in 2021 in the hopes of mitigating the introduction into the country. Most notably, Iraq implemented a targeted travel ban on India after the first detection of the Delta variant, from 27 April 2021 to 30 August 2021, and restricted entry for recreational activities by implementing a tourist travel restriction that was in place from 15 February 2021 to April 2022.^4,18,19^ However, despite these travel restrictions, Iraq experienced a large outbreak of the Delta variant during the second half of 2021 similar to other countries, followed by an outbreak of the Omicron variant in the first quarter of 2022. Though the travel restrictions did not prevent a surge in cases, it remains uncertain whether these measures delayed the introduction of the VOCs or the establishment of community transmission within Iraq. Additionally, even if restrictions successfully prevented the introduction of the Delta variant from India, it is unclear which alternative countries served as primary sources. This is further complicated by the limited sampling in the region, leaving unanswered questions about the role of regional connectivity in the spread of the virus in the region, as Iraq is highly socio-politically and economically connected to its neighbors.

To address these questions, we generated 535 whole-genome sequences of SARS-CoV-2 from clinical samples collected in Duhok, Iraq, between June 2021 and March 2022, and reconstructed the spatiotemporal spread of the virus using BEAST.^20^ Our analysis revealed that Delta was introduced in early February 2021, at least three months before the implementation of the targeted travel ban, while the first introduction of Omicron occurred in late November 2021 despite the ongoing tourist travel restriction. Both variants rapidly established community transmission, with 75-80% of new cases emerging from lineages persisting within Iraq after two months. These findings indicate that travel restrictions had limited efficacy in both preventing the introduction of the virus and successfully establishing community transmission within Iraq. We observed that this pattern was consistent on a global scale, with variants typically introduced before travel restrictions were implemented. Taken together, these findings highlight that these types of restrictions are ineffective and comprehensive genomic surveillance across multiple countries is essential for early detection and containment of emerging variants of concern.

## Results

### Improved genomic sampling in Iraq reveals dominant sublineages of SARS-CoV-2 variants in the wider region

To address the lack of sufficient genomic sampling in Iraq, we generated a total of 535 viral genomes between June 2021 and March 2022 using clinical samples from hospitalized patients in Duhok, Iraq (**Fig. 1A**). Our study period captured 343 genomes during the emergence and subsequent decline of the Delta variant, followed by 180 genomes of the Omicron variant during its emergence and subsequent decline (**Fig. 1B**).

**Figure 1.**
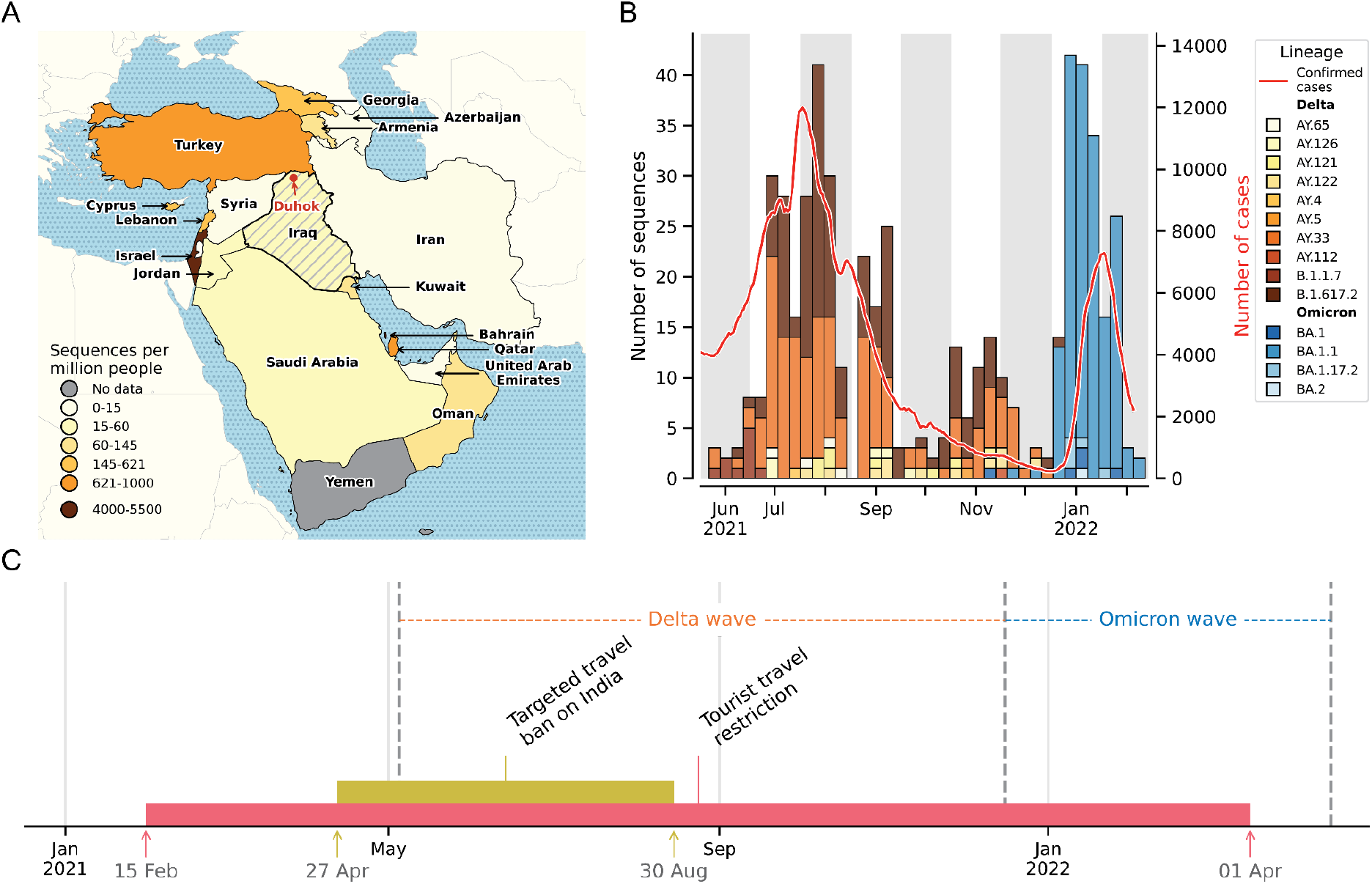
Improved sampling of SARS-CoV-2 genomes in Iraq reveals dominant sublineages of VOCs. (**A**) Geographic distribution of sequences collected per million residents in 2021 across Iraq and its neighboring countries, with Duhok in Iraq highlighted by a red dot. (**B**) Number of sequences collected by month (bar graph, left axis), colored by lineage classifications and the number of COVID-19 cases (7-day rolling average) (line graph, right axis). (**C**) Timeline of travel restrictions enacted by the Iraqi government.

In our dataset, the earliest sample of the Delta variant in Iraq was collected on 8 June 2021. The Delta variant drove the third surge in cases in the country from May to December 2021 (**Fig. 1B**). We found that the ancestral B.1.617.2 lineage and sublineage AY.33 were predominant, with several other sublineages co-circulating at lower prevalences (**Fig. 1B**). As the surge in cases due to the Delta variant subsided, cases due to the Omicron variant began to surge in Iraq, with our earliest sample dating back to 27 November 2021. The Omicron variant led to the fourth surge in cases from January to March 2022, with BA.1.1 as the predominant lineage (**Fig. 1B**). In response to the spread of the VOCs, Iraq imposed travel restrictions to delay the introduction into the country (**Fig. 1C**). However, the surge in cases indicates that these travel measures were ineffective in blocking the introduction of the variants. It remains unclear whether the restrictions delayed the arrival of VOCs or the onset of community transmission within Iraq, or how the source countries of viral introductions shifted over time in response to travel restrictions.

### Travel restrictions were ineffective in delaying the introductions of SARS-CoV-2 variants

To estimate the impact of the travel restrictions on the introduction of the Delta and Omicron variants in Iraq, we reconstructed the spatiotemporal spread of the virus using a discrete Bayesian phylogeographic analysis as implemented in BEAST^20^. We used the Delta variant genomes to evaluate the targeted travel ban between 27 April 2021 and 30 August 2021 (**Fig. 1C**).^4,18^ We found that for the Delta variant, Southern and Western Asia were the major sources of introductions into Iraq, with very few introductions coming from other parts of the world. We estimated a total of 12 (95% HPD: [6, 16]) and 7 (95% HPD: [2, 15]) introductions from Southern and Western Asia, respectively (**Fig. 2A**). This is in line with previous studies showing that the Delta variant emerged in India in mid-October and spread globally by early May 2021.^14^ The high number of introductions from Western Asia strongly suggests that introductions of the Delta variant reached Iraq not only through direct travel from India but also via neighboring Western Asian countries.

**Figure 2.**
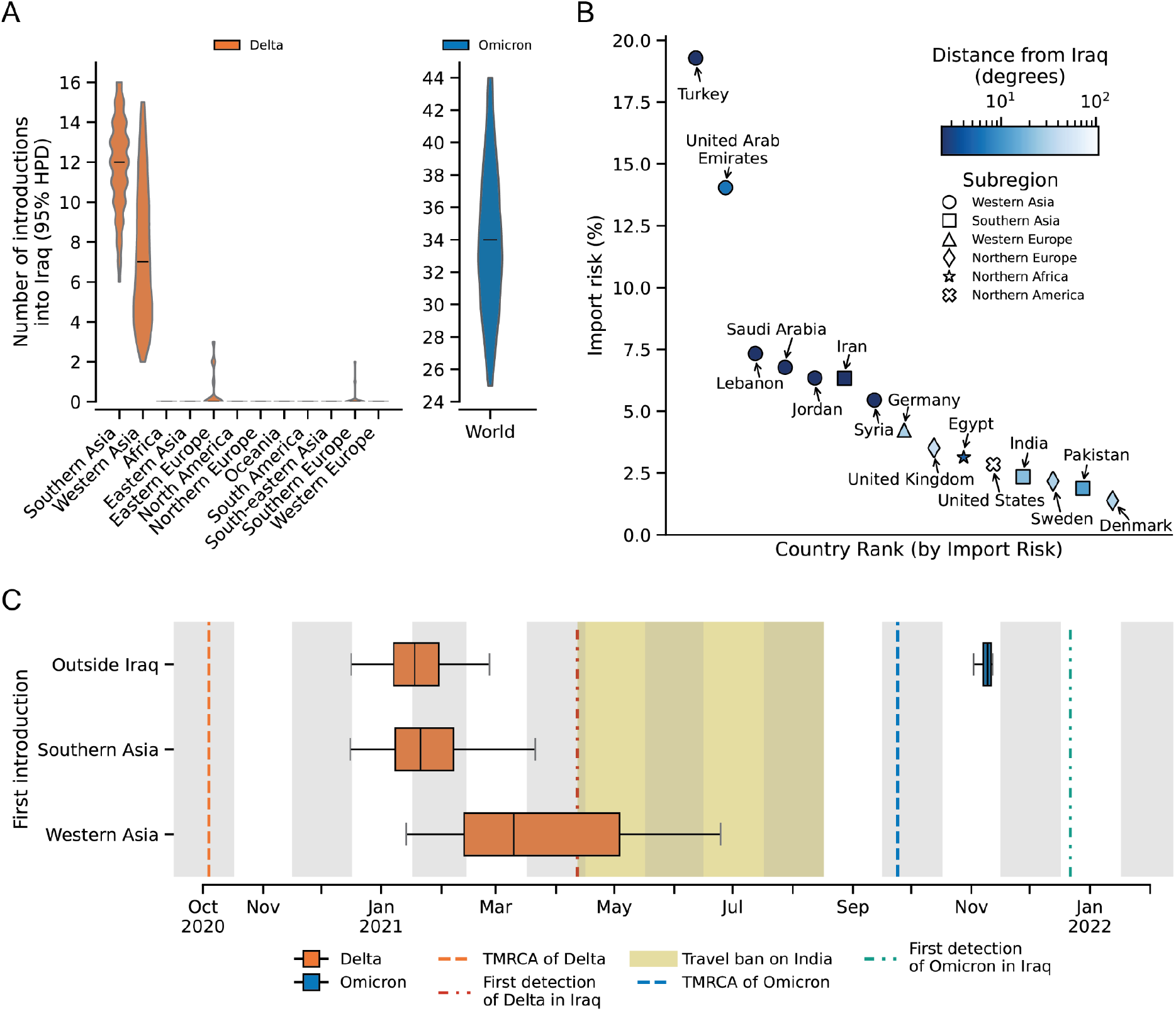
Impact of travel restrictions on the introduction of the Delta and Omicron variants into Iraq. (**A**) Estimated number of introductions of the Delta (orange) and Omicron (blue) variants into Iraq (95% HPD shown as a violin plot with the median indicated by the bar). **(B)** Distribution of estimated import risk percentages to Iraq from the top 15 contributing countries. **(C)** Estimated first introduction of the Delta and Omicron variants into Iraq from Western and Southern Asia, and the global region outside of Iraq, respectively.

To assess the efficacy of the tourist travel restriction imposed between 15 February 2021 and 1 April 2022, we analyzed the Omicron wave in Iraq (**Fig. 1C**).^19,21^ Given the rapid spread and potential sampling bias of the Omicron variant, we used phylogenetic analysis to estimate the total number of introductions into Iraq and an import risk model based on total air passenger volumes from October 2021 to April 2022 to estimate the risk of importation of the Omicron variant from other countries into Iraq (see, Methods). Our phylogeographic analysis estimated a median of 34 (95% HPD: [25, 44]) introductions of the Omicron variant into Iraq from the rest of the world (**Fig. 2A**). The import risk model estimated that Turkey and the United Arab Emirates posed the greatest importation risk, contributing ∼19% and ∼14% of the total incoming risk, respectively (**Fig. 2B**). Regional neighbors including Saudi Arabia, Iran, Lebanon, and Jordan, collectively accounted for nearly 27% of the importation risk (**Fig. 2B**). Notably, countries farther away from Iraq, such as the United Kingdom, United States, and Germany, still ranked among the top fifteen due to high travel volumes (**Fig. 2B**). These findings align with previous research showing that the United States, the United Kingdom, and Turkey were significant sources of Omicron BA.1 exports into other parts of the world.^5^

Although we have estimated that a high number of introductions have occurred, their timing and duration of cryptic spread before detection remain uncertain. To estimate the lag between the introduction and detection of each variant, we calculated the difference between the earliest introduction and the initial report of detection. Overall, we observed a lag of at least one month between the first introduction and first detection of SARS-CoV-2 variants in Iraq. For the Delta variant, the targeted travel ban was implemented one month after the variant was already spreading cryptically within Iraq (**Fig. 2C**). For the Omicron variant, the tourist travel restriction was ineffective at preventing the introduction of the variant (**Fig. 2C**). We estimate that the first introduction of the Delta variant from Southern Asia occurred in early February 2021 (95% HPD: [31 December 2020, 5 April 2021]) with the first introduction from Western Asia in late March (95% HPD: [28 January 2021, 9 July 2021]) (**Fig. 2C**). These introductions were at least one month before the first detection of the variant in Iraq, and approximately three months after its estimated emergence in India in October 2020.^14,18,22^ The first introduction of the Omicron variant in our dataset occurred in late November 2021 (95% HPD: [17 November 2021, 26 November 2021]), which is approximately one month after it was first detected in South Africa and one month before its detection in Iraq (**Fig. 2C**).^16,23^ Taken together, these findings show that travel restrictions are unlikely to substantially delay the introduction of rapidly spreading SARS-CoV-2 variants.

### Travel restrictions failed to prevent the rapid establishment of community transmission of the Delta and Omicron variants

Although we found that travel restrictions failed to delay the introduction of variants into Iraq, the extent to which they curbed introductions after implementation is unknown. We estimated that most introductions of the Delta variant from Southern Asia and Western Asia into Iraq occurred after the targeted travel ban was enacted (27 April 2021 to 30 August 2021). Moreover, for both the Delta and Omicron variants, the majority of sampled introductions occurred during periods of observed exponential epidemic growth (**Fig. 3A-B**). These observations further underscore that travel restrictions are unlikely to substantially delay the introduction of a rapidly spreading VOC.

**Figure 3.**
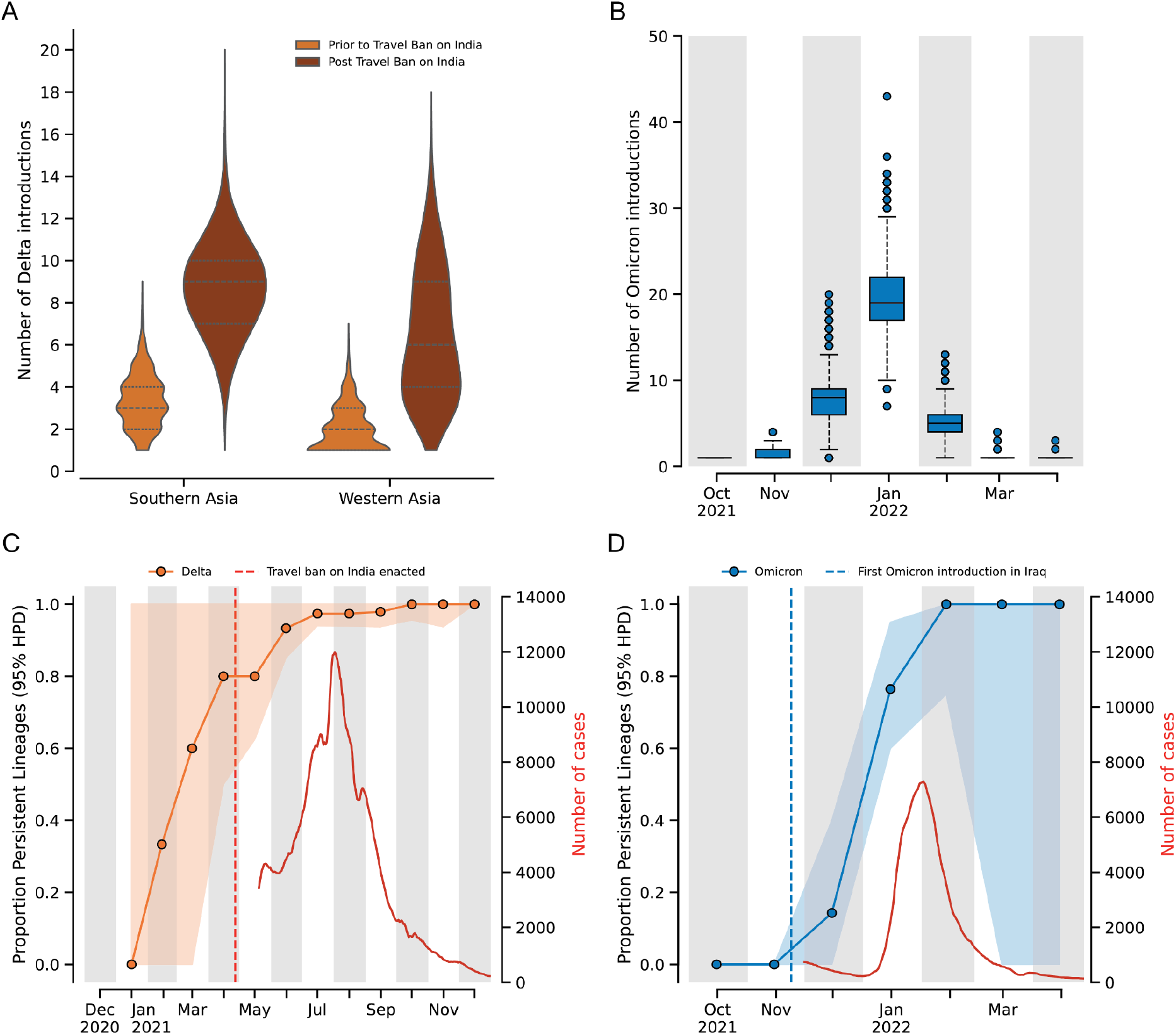
Dynamics of Delta and Omicron variant introductions and persistence of the variants within Iraq. **(A)** Violin plots showing the number of Delta variant introductions into Southern and Western Asia before and after the implementation of travel restrictions on travel to and from India. **(B)** Boxplots show the number of introductions of the Omicron variant over time from October 2021 to March 2022. Temporal dynamics of the proportion of persistent lineages (95% HPD) for (**C**) Delta and (**D**) Omicron variants.

To determine if the surge in cases due to the Delta and Omicron variants was driven by new introductions from outside Iraq or transmission from locally-established lineages, we estimated the proportion of lineages that were due to persistent transmission within Iraq over one-month intervals using PersistenceSummarizer available in BEAST. For each interval, the proportion of lineages arising from introductions predating that interval was calculated across the posterior distribution of phylogenetic trees (see, Methods). This approach enabled the quantification of the relative contributions of persistent local transmission versus new introductions from outside the country towards ongoing transmission within each interval. For the Delta variant, we found that a median of 80% (95% HPD: [50%,100%]) of transmission was due to introductions that occurred before the implementation of the targeted travel ban (**Fig. 3C**). Similarly, within two months of the first introduction of the Omicron variant into Iraq, a median of 76% (95% HPD: [60%, 95%]) of transmission resulted from already established local lineages (**Fig. 3D**). These findings demonstrate that the early introductions led to the rapid establishment of community transmission of VOCs within Iraq despite the travel restrictions. The rapid establishment of persistent lineages of both variants (median >75% for both) suggests that travel restrictions were ineffective in preventing community transmission within Iraq, with most introductions seeding local transmission chains.

### Global-scale analysis reveals travel restrictions were ineffective in preventing cryptic community transmission of SARS-CoV-2 variants

In previous sections, we demonstrated that the travel restrictions were implemented too late to prevent initial introductions into Iraq, while subsequent introductions were not only unimpeded by the restrictions but actually increased after their implementation. Furthermore, the early introductions into Iraq established community transmission of VOCs within Iraq, rendering the travel restrictions ineffective. To evaluate this pattern at a global scale, we analyzed data from Tegally et al. 2023^5^. We compared the estimated first introduction times of variants into countries worldwide with the timing of travel restrictions each country implemented to delay variant introduction. This comparison allowed us to calculate the lag between variant introduction and implementation of travel restrictions worldwide (Supplementary Data). To account for high variability in genomic sampling and travel between countries, we analyzed countries characterized by high passenger volumes from the variant’s country of first detection (greater than the median passenger volume) and robust genomic surveillance systems (greater than the median genomic sampling rate), as shown in the top-right quadrant of **Fig. 4A** and **4B**. Despite robust genomic surveillance infrastructure in these countries, we found that the median interval between first inferred Delta introduction and implementation of travel restrictions was greater than 1 month later across all countries investigated (**Fig. 4C**). We observe a similar pattern for Omicron, with introductions occurring a median of 10 days before travel restrictions were implemented (**Fig. 4D**). The outliers in Fig. 4C and 4D including The Netherlands, Canada, and Mauritius, are the result of a lack of sampling of early cases since these countries reported detections of these variants before widespread global detection.^6,24,25^ Notably, even countries such as the island continent Australia, with near-complete closure of international borders, only delayed the introduction of the Delta variant by a median of 49 days without preventing its spread within the country.^26^ This analysis reveals that the limited effectiveness of travel restrictions on mitigating the introductions of novel SARS-CoV-2 variants, as seen in Iraq, was similarly observed at a global scale. Collectively, these findings emphasize that travel restrictions are often implemented too late to prevent the introduction of VOCs and subsequent transmission within the region.

**Figure 4.**
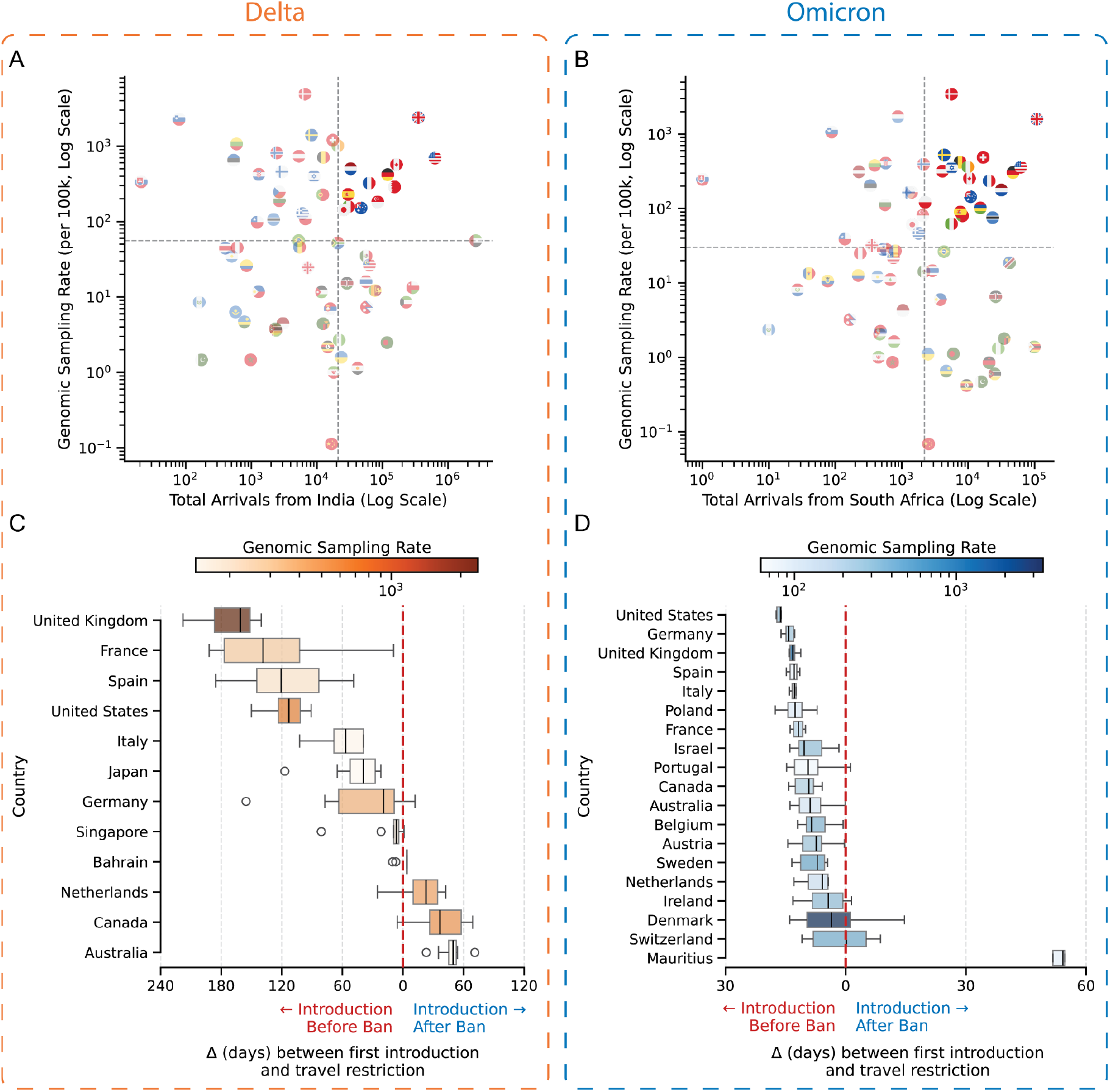
Impact of travel restrictions on introduction of the Delta and Omicron variants on a global scale. Scatter plot showing genomic sampling rate and the number of total arrivals from (**A)** India during the emergence of the Delta variant and (**B**) South Africa during the emergence of the Omicron variant. Each point represents a country, indicated by its flag. Boxplots showing the delay (in days) between the first detected introduction of the variant and the implementation of travel restrictions for (**C**) Delta and (**D**) Omicron variants. Negative values (left of the dashed vertical line) indicate introductions prior to targeted travel bans; positive values (right) indicate introductions after the bans. The color of the box plot indicates the genomic sampling rate.

## Discussion

In this study, we used phylogeographic analyses to investigate the effectiveness of targeted travel bans and tourist travel restrictions implemented in Iraq during the third and fourth COVID-19 waves. We found that the Delta variant was introduced three months before the targeted travel ban was implemented, with community transmission established approximately two months prior to the restrictions. Similarly, the Omicron variant was introduced one month before it was first detected and had begun circulating within the country despite the tourist travel restriction. These findings suggest that the targeted travel ban was implemented far too late, relative to the emergence and cryptic transmission of Delta, to effectively mitigate the spread of the virus into Iraq. It also supports that the tourist travel restriction is ineffective against a rapidly spreading variant such as Omicron. We also observed that early introductions predominantly originated within Western Asia, indicating that most of the introductions of the virus into Iraq came from neighboring countries. Thus, in the case of a rapidly dispersing pathogen, targeted travel bans are ineffective due to potential indirect introductions through intermediate countries. We also show that tourist travel restrictions are ineffective at preventing the introduction of a VOC.

Our findings reveal fundamental limitations in travel restrictions as a pandemic control measure. The consistent pattern of delayed detection, with variants establishing community transmission weeks to months before implementation of travel restrictions, demonstrates that such measures are often too late to meaningfully impact the rapid spread of a pathogen.^5,27^ The dominance of indirect introductions of the virus through intermediate countries over primary source countries in global variant exportation further undermines the effectiveness of travel restrictions.^5^ These results highlight that pandemic preparedness strategies must shift from reactive border controls toward comprehensive, internationally coordinated genomic surveillance systems capable of detecting emerging variants in real-time.

Our findings should be contextualized by our limited sample size, which represents less than 0.01% of cases within the study period. Therefore, the number of single introductions and the size of the transmission chains that we report may be severely underestimated.^2^ Moreover, regional introductions from neighboring and regional West Asian countries are likely to be underestimated due to undersampling of those regions.^11^ Our sampling was concentrated in Duhok, a city in northern Iraq close to the border with Turkey and Syria, and may not be representative of population-level transmission dynamics in the rest of the country. For example, regions near the border may experience more introductions from neighboring countries due to shorter land-based travel. However, it is highly likely that earlier introductions of these variants and community transmission occurred outside of Duhok.

Scientific evidence from COVID-19 demonstrates that rapid genomic sequencing and real-time international data sharing are our most effective tools against emerging VOCs.^2,3,7–9^ Within days of Omicron’s identification in South Africa, global sequencing networks had characterized its mutations and transmission patterns, information that proved far more valuable than the travel restrictions that followed.^16^ Research shows VOCs typically spread internationally before detection, making border controls largely ineffective while potentially discouraging countries from transparent reporting due to economic consequences.^28–31^ Public health responses should prioritize non-pharmaceutical measures such as stay-at-home orders, mask mandates, and limits on gatherings alongside vaccination, specifically restricting gatherings to fewer than 10 people, closing high-exposure businesses, and closing schools and universities, which have been shown to work.^28–31^

## Methods

### Sample collection

SARS-CoV-2 samples were collected from routine diagnostic qPCR tests performed by the University of Duhok COVID-19 center, Duhok, Iraq. To ensure a high success rate, we only sequenced positive qPCR samples with a Ct value less than 30. In total, we collected and sequenced 535 SARS-CoV-2 samples. The samples were collected from June 2021 until February 2022, spanning the beginning of the B.1.1.7 wave, the entire Delta wave (June to December 2021), and the beginning of the Omicron wave. All sequence data generated for this study have been made publicly available on GISAID (GISAID accession IDs are available on https://github.com/andersen-lab/HCoV-19-Genomics)^32^

### SARS-CoV-2 whole-genome sequencing

Samples were shipped to the Andersen lab at Scripps Research on dry ice, and sequenced using an amplicon-based sequencing assay using overlapping ∼250nt amplicons.^33^ Briefly, viral RNA was extracted using the Mag-Bind Viral DNA/RNA kit (Omega Bio-tek) according to the manufacturer’s instructions. SARS-CoV-2 RNA (2 µL) was reverse transcribed with SuperScript IV VILO (ThermoFisher Scientific). The virus cDNA was amplified in two multiplexed PCR reactions using Q5 DNA High-fidelity Polymerase (New England Biolabs). Following an AMPureXP bead (Beckman Coulter) purification of the combined PCR products, the samples were barcoded in another PCR reaction using barcoded Illumina adapters as primers. The libraries were again purified with AMPureXP beads and quantified using the Qubit High Sensitivity DNA assay kit (Invitrogen) and Tapestation D5000 tape (Agilent). The individual libraries were then normalized and pooled in equimolar amounts at 1.5 nM. The 1.5 nM library pool was sequenced using a NovaSeq 6000 SP Reagent Kit v1.5 (300 cycles). Consensus sequences were assembled using an in-house Snakemake pipeline with bwa-mem and iVar v1.2.2.^34–36^

### Downsampling strategies and dataset curation

To generate a downsampled dataset representative of the global diversity of the variants for phylogenetic analyses, we first downloaded all publicly available sequences of the lineages B.1.617.2, AY.4, AY.5, AY.33, AY.112, AY.121, AY.122, and AY.126 until May 2022 from GISAID. Sequences with low coverage (>1% N’s and >0.5% unique amino acid mutations) or with incomplete collection dates were excluded. Sequences were randomly subsampled proportional to the weekly number of new positive COVID-19 cases in each country. The number of new COVID cases per country was obtained from the COVID-19 dataset generated by Our World in Data.^37^

The downsampled sequence dataset was aligned to the reference genome Hu-1 (GenBank accession number: MN908947.3) using MAFFTv7.505, and the 5’ and 3’ untranslated regions were trimmed, and all problematic sites in the alignment were masked.^38,39^ The masked, aligned dataset was then used to reconstruct a maximum likelihood phylogenetic tree with IQTREE-2 using ModelFinder and used TreeTime to generate a root-to-tip plot.^40–42^

Finally, sequences clustering within a genetically divergent subclade of the B.1.617.2 lineage were removed. The final downsampled and curated dataset contained 1,152 sequences. Pangolin^43^ was used to assign a lineage to each sequence in the alignment.^43^ A list of GISAID IDs for our final downsampled sequence dataset can be found using the following GISAID EPI_SET ID: EPI_SET_221201da.^44^

For the phylogenetic analysis of the Omicron variant, we downsampled BA.1, BA.1.1, and BA.1.17.2 variants from September 2021 to May 2022 from GISAID. Sequences with low coverage (>1% N’s and >0.5% unique amino acid mutations) or incomplete collection dates, or early sequences not part of the wave.

For Omicron, to minimize bias from under-reported cases, we employed uniform sampling: 2 sequences per country per week, with neighboring countries (Armenia, Azerbaijan, Bahrain, Cyprus, Iran, Israel, Jordan, Kuwait, Lebanon, Oman, Qatar, Saudi Arabia, Syria, Turkey, United Arab Emirates, Yemen) up-weighted by a factor of 2.

We reconstructed a maximum likelihood phylogenetic tree from the final dataset of 1,711 sequences. Pangolin was used to assign a lineage to each sequence in the alignment.^43^ GISAID IDs for the downsampled dataset are available via GISAID EPI_SET ID: EPI_SET_250114rz^45^, plus four additional sequences (EPI_ISL_1227819, EPI_ISL_12604523, EPI_ISL_12604439, EPI_ISL_12278224) can be found here: https://github.com/andersen-lab/HCoV-19-Genomic

### Phylogenetic analyses

BEAST v1.10.5^20^ was used to reconstruct a time-scaled phylogeny under a HKY substitution model with gamma-distributed rate variation among sites, a strict clock with a lognormal prior, and a flexible Skygrid coalescent prior with grid points every two weeks.^20^ For the phylogenetic analysis of the Omicron variant, we specified standard transition kernels on all parameters except for the parameters of the skygrid coalescent model. For these parameters, we used Hamiltonian Monte Carlo transition kernels to improve sampling efficiency. For the phylogenetic analysis of the Delta variant, we specified standard transition kernels on all parameters. We performed inference under the fully specified model using Markov Chain Monte Carlo (MCMC) sampling, the BEAGLE v4 library for parallelization. The MCMC chain was run for 200 million iterations with trees sampled every 10,000 steps.^20^ Tracer v1.7.2^46^ was used to assess the convergence (Effective Sample Size >100) of the chain after removing a 10% burn-in. We used TreeAnnotator 1.10 to obtain the maximum clade credibility phylogenetic tree and used the Baltic library to visualize the tree.^47,48^

To reconstruct the phylogeographic history for the Delta variant, we performed an asymmetric discrete state analysis using an empirical set of trees to reconstruct transmission dynamics between Iraq and geographic regions (as defined by the UN) by summarizing the start and end location of Markov jumps across the full posterior of all trees using TreeMarkovJumpHistoryAnalyzer.^48,49^

### Import Risk Model

Monthly air passenger volume data from October 2021 to April 2022 were obtained from Bluedot.^50^ The import risk from a region is defined as the proportion of total passenger volume coming into Iraq from a given region during the study period.

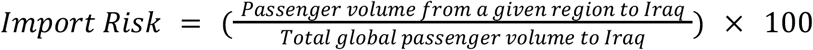

The import risk model assumes that travel intensity serves as a proxy for the risk of introduction of a variant from each respective region. Given the rapid global dissemination characteristics of the Omicron variant, the analysis focused on passenger volume patterns rather than global connectivity patterns.

### Persistence summarizer

To determine the number of unique introductions and the occurrence of community transmission, we summarized the number of unique circulating lineages and the number of descendants in a given time frame across the complete posterior distribution of trees using PersistenceSummarizer.^51^

## Data availability

The data for our analyses can be found at https://github.com/andersen-lab/paper_2025_SARS-CoV-2_Cryptic_Spread. All sequence data generated were made available via the GISAID database (GISAID EPI_SET ID: EPI_SET_221201da, EPI_SET_250114rz, and four additional sequences EPI_ISL_1227819, EPI_ISL_12604523, EPI_ISL_12604439, EPI_ISL_12278224). Air travel data can be requested for release from Bluedot.

## Acknowledgements

The research leading to these results has received funding from the National Institutes of Health grants U19AI135995 and U01AI151812.

## Notes

### Competing Interest Statement

The authors have declared no competing interest.

